# High infection attack rates of SARS-CoV-2 in Dutch households revealed by dense sampling

**DOI:** 10.1101/2021.01.26.21250512

**Authors:** Daphne F.M. Reukers, Michiel van Boven, Adam Meijer, Nynke Rots, Chantal Reusken, Inge Roof, Arianne B. van Gageldonk-Lafeber, Wim van der Hoek, Susan van den Hof

## Abstract

**Background:** Indoor environments are considered a main setting for transmission of SARS-CoV-2. Households in particular present a close-contact environment with high probability of transmission between persons of different ages and with different roles in society.

**Methods:** Complete households with a laboratory-confirmed SARS-CoV-2 positive case in the Netherlands (March-May 2020) were included. At least three home visits were performed during 4-6 week of follow-up, collecting naso- and oropharyngeal swabs, oral fluid, faeces and blood samples for molecular and serological analyses of all household members. Symptoms were recorded from two weeks before the first visit up to the last visit. Secondary attack rates (SAR) were estimated with logistic regression. A transmission model was used to assess transmission routes in the household.

**Results:** A total of 55 households with 187 household contacts were included. In 17 households no transmission took place, and in 11 households all persons were infected. Estimated SARs were high, ranging from 35% (95%CI: 24%-46%) in children to 51% (95%CI: 39%-63%) in adults. Estimated transmission rates in the household were high, with reduced susceptibility of children compared to adolescents and adults (0.67; 95%CI: 0.40-1.1).

**Conclusion:** Estimated SARs were higher than reported in earlier household studies, presumably owing to a dense sampling protocol. Children were shown to be less susceptible than adults, but the estimated SAR in children was still high. Our results reinforce the role of households as main multiplier of SARS-CoV-2 infection in the population.

**Key points:** We analyze data from a SARS-CoV-2 household study and find higher secondary attack rates than reported earlier. We argue that this is due to a dense sampling strategy that includes sampling at multiple time points and of multiple anatomical sites.

## Introduction

The first case of the coronavirus disease (COVID-19) emerged in Wuhan in December 2019 [1]. Starting with an outbreak of pneumonia of unknown etiology, the causative agent severe acute respiratory syndrome coronavirus 2 (SARS-CoV-2) was identified early January 2020 [2]. Since then, the virus has spread rapidly across the world [3].

Evidence from case and cluster reports shows that SARS-CoV-2 is largely spread through respiratory droplets from infected persons, with proper distance and indoor air ventilation being significant factors reducing the risk of transmission [4]. Therefore, social distancing measures are important to reduce transmission, and most countries have instated strategies based on this premise. In the Netherlands, the first COVID-19 case was detected on February 27 [5]. In March, the Dutch government mandated a partial lockdown, characterized by social distancing, self-quarantine and self-isolation orders, closing of schools, bars, and restaurants, and urging people to work from home [6]. These measures generally increased the time spent at home. As household members live in close contact it is difficult to attain a proper physical distance after a COVID-19 diagnosis of a household contact. In combination with evidence that a sizeable fraction of transmission events occur pre-symptomatically, the household constitutes a high risk setting for SARS-CoV-2 transmission [7].

The secondary attack rate (SAR) of SARS-CoV-2 infection among household contacts is a useful measure to gauge the risk of transmission in this close-contact setting. It provides insight in the susceptibility of contacts and infectiousness of cases given certain characteristics, such as age, gender, household size, and severity of infection. Household studies performed in the first six months of the pandemic, mostly in China, found a relatively high household SAR of 15-22% [8]. In most countries, paediatric patients are underrepresented in the statistics of the COVID-19 outbreak and children usually exhibit mild symptoms [9, 10]. If children have lower susceptibility or infectiousness, this can have important implications for strategies to curb the spread of SARS-CoV-2. Previously household studies observed that the SAR was significantly higher for adult contacts compared to child contacts [11]. However, most studies only tested household contacts with COVID-19 related symptoms, relied on RT-PCR in nasopharyngeal swabs only, and did not perform any follow-up sampling. These studies may have missed mild, pre- or asymptomatic cases, especially in children [12, 13]. In the present study all household contacts were tested as soon as possible after a laboratory-confirmed infection in the household was established, and subsequently followed-up for 4 to 6 weeks. A dense sampling strategy was employed that included sampling from various anatomical sites while using multiple molecular and serological diagnostic methods to establish infection. This increases the chance of detecting every SARS-CoV-2 infected household contact and of determining transmission routes, including asymptomatic transmission, as accurately as possible [8]. Main aims of this study were to estimate secondary attack rates and to determine factors that impact susceptibility and infectiousness, with a specific focus on age of household contacts.

## Methods

This study is an update of a generic stand-by protocol drafted in 2006 to quickly initiate scientific research in the case of an outbreak of an emerging pathogen [14]. The generic protocol was tailored to the current COVID-19 pandemic with input from the WHO First Few Hundred protocol [15]. The generic and adapted study protocols were approved by the Medical-Ethical Review Committee of the University Medical Center Utrecht (NL13529.041.06). A prospective cohort study was performed following households where one household member was tested positive for SARS-CoV-2 in the period March 24-April 6 2020 (one household was included later on May 24).

### Population

Any person 18 years and older testing positive for SARS-CoV-2 who had at least one child in their household below the age of 18 and consented to be contacted for scientific research were reported by the Public Health Service of the region Utrecht. We contacted this person (i.e. the index case) to request enrolment of the entire household in this study. Every household contact (persons living in the same house as the index patient) was to be enrolled in the study, except for contacts below the age of one year. Households were excluded if one or more of the household contacts did not want to participate in the study upfront, as in that case it would not be possible to fully determine household transmission patterns.

### Data collection

Two research nurses performed the first home visit within 24 hours after inclusion to collect the informed consent forms and the first samples from all participants (see Table 1 for schedule of sample collection). Household contacts completed a questionnaire to collect demographic characteristics, medical history, travel history, anti-viral drug use, symptoms, symptom onset and hospital admission. Participants reported whether they had symptoms in the 2 weeks prior to the first visit. After the first visit, they filled in a symptoms diary for 2 weeks. A second visit was included at 2-3 weeks post-inclusion, and at the last home visit at 4-6 weeks post-inclusion, participants reported whether they had developed symptoms in the weeks between the second and third home visit. We defined three age strata: adults 18 years of age or older, adolescents 12 to 17 years of age (corresponding to secondary school age) and children 1 to 11 years of age (corresponding to day care and primary school age).

**Table 1.** Schedule of administering questionnaires, symptom diaries and home visits for sampling by a research nurse.

|  | Day | 1 | 2 | 3 | 4 | 5 | 6 | 7 | 8 | 9 | 10 | 11 | 12 | 13 | 14 | 15 (range<br>14-21) | 35 (range<br>28-42) |
| --- | --- | --- | --- | --- | --- | --- | --- | --- | --- | --- | --- | --- | --- | --- | --- | --- | --- |
| **Basic questionnaire** |  | x |  |  |  |  |  |  |  |  |  |  |  |  |  |  |  |
| **Retrospective symptom questionnaire<sup>1</sup>** |  | x |  |  |  |  |  |  |  |  |  |  |  |  |  |  | x |
| **Symptom diary (on that specific day)** |  |  | x | x | x | x | x | x | x | x | x | x | x | x | x |  |  |
| **Serum<sup>2</sup>** |  | x |  |  |  |  |  |  |  |  |  |  |  |  |  | x | x |
| **Naso- and oropharyngeal swab<sup>3</sup>** |  | x |  | (x) |  |  | (x) |  |  | (x) |  |  | (x) |  |  | x |  |
| **Oral fluid** |  | x |  |  |  |  |  |  |  |  |  |  |  |  |  | x | x |
| **Feces<sup>4</sup>** |  | x |  |  |  |  |  |  |  |  |  |  |  |  |  | x | x |

<sup>1</sup> This questionnaire included questions on symptoms on the day of that home visit and symptoms in the 2 weeks prior to the first home visit or the symptoms between the last and second home visit.
<sup>2</sup> Participants ≥ 16 years of age: total of 18.5 ml. Participants with symptoms within 4 days prior to the first home visit, were asked to provide 3 extra tubes of 8 ml for additional cellular immunity assays. Participants <16 years: total of 13.5 ml. Participants also had the option of capillary finger blood collection (0.5 ml) instead of venous blood collection.
<sup>3</sup> Participants ≥ 16 years of age additionally had the option to get a naso- and oropharyngeal swab every 3 days (day 3, 6, 9 and 12). Participants of all ages without a previous positive SARS-CoV-2 result developing acute symptoms also received an additional naso- and oropharyngeal swab as soon as possible after developing these symptoms. A naso- and oropharyngeal swab was not collected for the index case at the first home visit, as these persons were already swabbed a few days before.
<sup>4</sup> Feces samples were not collected by the nurse visiting the household. Participants received a feces collection kit at the first home visit and were asked to collect feces within 3 days after the first, second and third home visit.

#### Molecular diagnostics and serological analysis

Total nucleic acid was extracted from the nasopharyngeal swab (NP), oropharyngeal swab (OP), oral fluid and faeces specimens using MagNApure 96 with total nucleic acid kit small volume and elution in 50 µl. RT-qPCR was performed on 5 µl extract using TaqMan® Fast Virus 1-Step Master Mix (Thermo Fisher) on Roche LC480II thermal cycler with SARS-like beta coronavirus (Sarbeco) specific E-gene primers and probe as described previously [16]. As no other Sarbeco viruses are currently detected in humans, a positive Sarbeco E-gene RT-qPCR is validly taken as positive for SARS-CoV-2. The results of the NP and OP swabs were combined to one result: upper respiratory tract (URT) negative (NP and OP negative) or positive (NP and/or OP positive). For detection of antibodies against SARS-CoV-2 we used the Wantai total Ig ELISA as described previously [17].

#### Classification of index and primary case

Laboratory-confirmed SARS-CoV-2 infection was defined as, either at least one positive PCR on any of the clinical samples taken during follow-up and/or detection of antibodies at any sampling timepoint. Every index case was by definition infected, as they had at least one positive PCR on an URT swab.

#### Symptoms and severity of COVID-19

The day of symptom onset as reported by the participant was set as the first day of illness. Participants were considered symptomatic if at least one of the following symptoms occurred at any timepoint: respiratory symptoms (including sore throat, cough, dyspnoea or other respiratory difficulties, rhinorrhoea), fever, chills, headache, anosmia or ageusia, muscle pain, joint ache, diarrhoea, nausea, vomiting, loss of appetite or fatigue. For household contacts, symptom onset occurring more than 2 weeks prior to the first day of illness or first positive test result of the index case were considered not related to SARS-CoV-2 transmission within the household. A differentiation was made between mild, moderate and severe COVID-19 based on self-reported symptoms or hospital admission [18]. We defined mild COVID-19 cases as laboratory confirmed cases showing any clinical symptoms. Moderate COVID-19 cases showed clinical signs of pneumonia, including dyspnea, and severe COVID-19 cases reported dyspnea and consulted a health professional (e.g. an emergency room) for their symptoms or reported having been admitted to the hospital for COVID-19.

#### Primary case

In every household, a primary case (the most likely first case of the household) was determined based on laboratory confirmation, symptom onset and travel history. A household contact was considered the primary case if they had a laboratory-confirmed SARS-CoV-2 infection with a symptom onset at least 2-14 days before the index case.

### Statistical analysis

#### Secondary attack rate

Household secondary attack rates (SARs) were estimated excluding the index case (i.e. the lab-confirmed person that led to inclusion of the household in the study), but including the primary case. This corresponds to common practice as reliable information on the primary case in the household often is lacking [19, 20]. To take clustered nature of the data into account, SARs were estimated with a logistic regression using generalized estimating equations (GEEs), with household as the unit of clustering and assuming an exchangeable correlation structure. Analyses were performed using three age strata as defined above, and using covariates sex, household size, and severity of infection of the index case. Model selection was based on the Quasi Information Criterion for small sample sizes (QICc). Analyses were carried out in R (version 3.6.0) using the geepack (version 1.5.1) and emmeans (version 1.3.1) packages.

#### Transmission model

Next to the estimates of the SAR we analysed the data using the final size distribution of a stochastic SEIR transmission model. In this model persons are classified as susceptible (S), infected but not yet infectious (E), infected and infectious (I), or recovered and immune (R). Appeal of these analyses are that the estimated parameters have a biological interpretation (susceptibility, infectiousness), that final-size distributions are invariant with respect to the latent-period distribution, and that different assumptions on the distribution of the infectious period can be incorporated [21, 22]. With respect to the contact process, we assumed frequency-dependent transmission as this mode of transmission is preferred over density-dependent transmission by information criteria (not shown) [23]. Time is rescaled in units of the infectious period, and we assumed a realistic variation in the infectious period, corresponding to an infectious period of 6-10 days. Here, because households were included only if an infected person was present, the final-size distributions needed to be conditioned on the presence of an infected index case if the index case was not also the primary case [22]. Such conditioning was applied for 17 households (Figure 1). Model selection was performed using LOOIC, a measure for predictive performance [23, 24]. Estimation was performed in a Bayesian setting using Hamiltonian Monte Carlo implemented in Stan (version 2.21.2) [25]. Details will be made available on our digital repository (https://github.com/mvboven/COVID-19-FFX).

**Figure 1.**
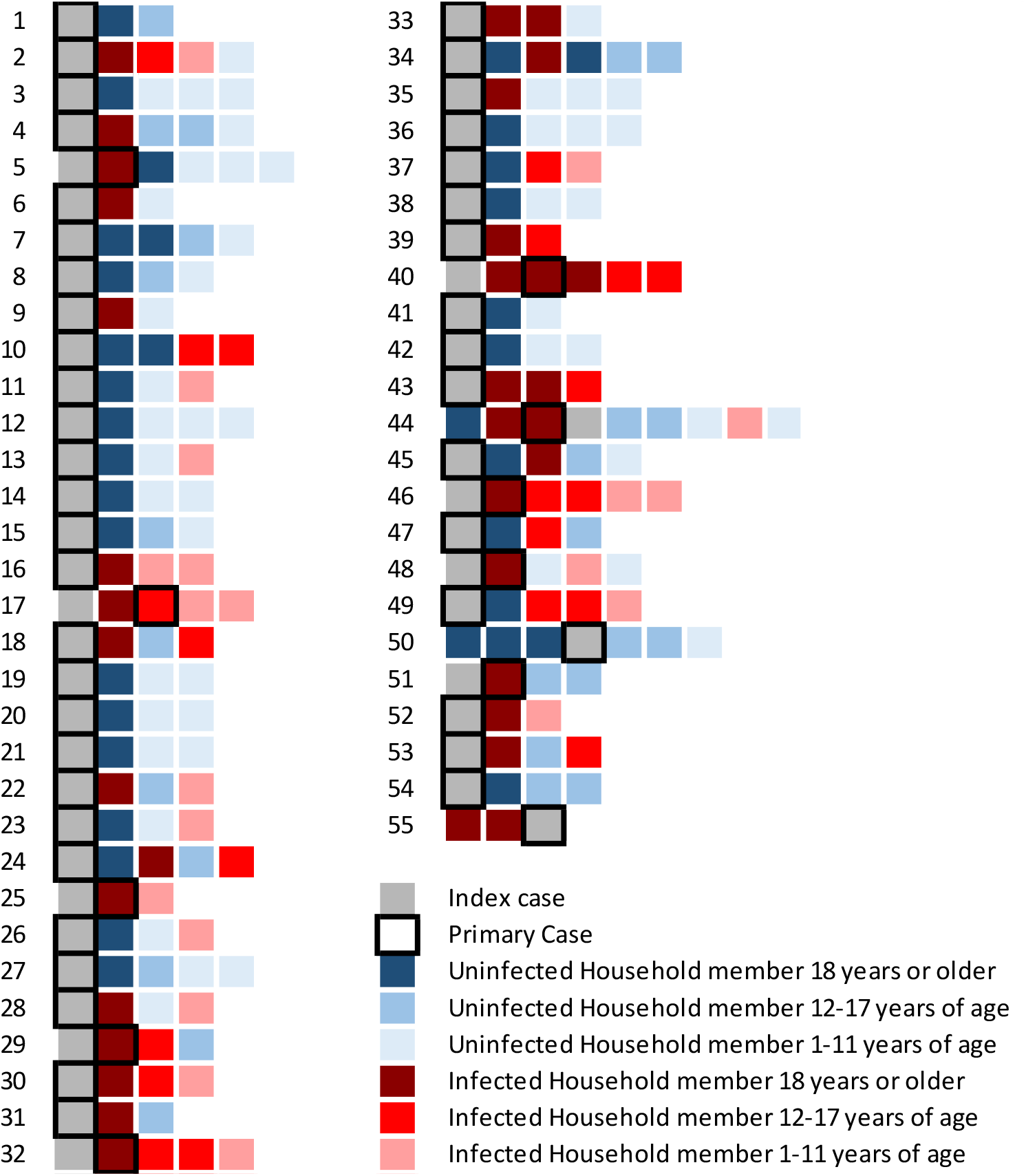
Overview of transmission of SARS-CoV-2 within households. Each row represents a household and each square a household member. The grey squares are the index cases and the most likely primary case is indicated by a black border. Blue squares indicate uninfected household members and red squares infected household members, with lighter colors indicating a younger age group. The squares are ordered by age, but the first two squares are always two spouses and the parents/guardians of the children.

### Ethics

The Medical Ethical Review Board of the University Medical Centre Utrecht reviewed and approved the study protocol (NL13529.041.06). All participants above the age of 12 gave written informed consent. Parents or guardians of participating children below the age of 16 gave written informed consent for participation, for children 12-16 both parents and children had to give consent.

## Results

### Descriptive analysis

Fifty-five households were included, with in total 242 participants of which 55 index cases and 187 household contacts (Table 2). Household size varied from three to nine persons (Figure 1). Index cases were predominantly female (n=40, 72.7%), and health care worker (n=41, 75.9%). Seven index cases were admitted to the hospital before or during participation in the study, and none of the other cases in the household required hospitalization. In 10 of the 55 households, the index case was observed not to be the primary case; in nine households this was determined to be another adult, and in one an adolescent contact. In 17 households, no transmission took place, and in 11 households every member got infected with SARS-CoV-2 (Figure 1). In children, fewer SARS-CoV-2 infections were found compared to adolescent and adult household contacts. In total, 51% of adults, 46% of adolescents, and 30% of children got infected. Children and adolescent household contact were less often symptomatic than adult contacts. Adults were also more likely to have a severe infection (37%) compared to adolescents (15%) and children (5%).

**Table 2.** Characteristics of the study population.

|  | All household members | Index cases | All household contacts | Adult household contacts<br>(≥ 18 years) | Adolescent household contacts<br>(12-17 years) | Children household contacts<br>(1-11 years) |
| --- | --- | --- | --- | --- | --- | --- |
|  | N=242<sup>1</sup> | N=55<sup>1</sup> | N=187<sup>1</sup> | N=71<sup>1</sup> | N=46<sup>1</sup> | N=70<sup>1</sup> |
| *N (%) unless stated otherwise* |  |  |  |  |  |  |
| **Age** (at first home visit) (*Mean (sd)*) | 27 (18.4) | 43 (9.0) | 22 (17.8) | 42 (12.8) | 14 (1.4) | 7 (3.0) |
| **Gender** (male) | 114 (47.1) | 15 (27.3) | 99 (52.9) | 44 (62.0) | 20 (43.5) | 35 (50.0) |
| **Education level** |  |  |  |  |  |  |
| Low |  | 1 (1.9) |  | 11 (15.9) |  |  |
| Medium |  | 17 (31.5) |  | 22 (31.9) |  |  |
| High |  | 36 (66.7) |  | 36 (52.2) |  |  |
| **Healthcare worker** |  | 41 (75.9) |  | 13 (19.1) |  |  |
| **Comorbidity** (one or more)<sup>2</sup> | 50 (21.1) | 20 (37.0) | 30 (16.5) | 16 (23.2) | 8 (18.2) | 6 (8.7) |
| **Symptomatic** |  |  |  |  |  |  |
| 2 weeks prior to first home visit | 158 (67.0) | 51 (94.4) | 107 (58.9) | 49 (71) | 24 (54.6) | 34 (49.3) |
| at first home visit | 126 (53.4) | 44 (81.5) | 82 (45.1) | 41 (59.4) | 16 (36.4) | 25 (36.2) |
| during follow-up | 126 (52.1) | 47 (85.5) | 79 (42.3) | 38 (53.5) | 17 (37.0) | 24 (34.3) |
| anytime during the study | 184 (78.0) | 54 (100) | 130 (71.4) | 56 (81.2) | 28 (63.6) | 46 (66.7) |
| **Hospital admission** (related to COVID-19) |  | 7 (12.7) |  |  |  |  |
| **Laboratory confirmed SARS-CoV-2 infection** | 133 (55.0) | 55 (100) | 78 (41.7) | 36 (50.7) | 21 (45.7) | 21 (30.0) |
| **Severity of infection** |  |  |  |  |  |  |
| Asymptomatic | 6 (4.7) | 0 | 6 (8.0) | 1 (2.9) | 3 (15.0) | 2 (10.0) |
| Mild | 73 (56.6) | 21 (38.9) | 52 (69.3) | 21 (60.0) | 14 (70.0) | 17 (85) |
| Moderate | 19 (14.7) | 11 (20.4) | 8 (10.7) | 6 (17.1) | 1 (5.0) | 1 (5.0) |
| Severe | 31 (24.0) | 22 (40.7) | 9 (12.0) | 7 (20.0) | 2 (10.0) | 0 |

<sup>1</sup> Six participants did not fill in the questionnaires. One index case, two adult contact, two adolescent contacts and one child contact. Information on education level, occupation, comorbidity, symptoms and severity is missing.
<sup>2</sup> Comorbidities include: asthma, COPD, chronic bronchitis or other chronic lung diseases, cardiovascular diseases, diabetes, immune disorders or received treatment causing immunocompromised state, chronic kidney, liver or neuromuscular disorders. Obesity, hypertension, hay fever, or other allergies are not included.

### Secondary attack rates

Overall estimated household secondary attack rate (SAR) was 43% (95%CI 33%-53%). In univariable analysis only age was significant at the 5% level (p=0.036), while sex (p=0.11), household size (p=0.64), being a healthcare worker (p=0.28) and severity of infection of the index case (p=0.30) were all not significantly associated with the outcome (p>0.10). In a multivariable analysis that included sex and age group (child, adolescent, adults), being a child was strongly associated with decreased probability of infection (p=0.006), while there was marginal evidence that female sex was associated with increased probability of infection (p=0.053). The univariable model with age was the preferred model based on QICc. Estimates of the SARs of this model show that SAR is lowest in children (35%, 95%CI: 24%-46%), higher in adolescents (0.41, 95%CI: 27%-56%), and highest in adults (51%, 95%CI: 39%-63%).

### Household transmission

Building on the results of the SAR estimates, we analyzed transmission models that differed with respect to assumptions on the susceptibility and transmissibility of age groups (children, adolescents, adults). Table 3 shows the results. In the unstructured model the transmission rate was estimated at 1.2 (unit: per infectious period). Given our assumption on frequency-dependent transmission this implies that the probability of direct transmission from an infected to an uninfected person (i.e. without taking indirect transmission via intermediate persons into account) in a household of four persons would be 1-exp[-1.2/4]=0.26. Overall, differences between models were modest, and the data did not allow estimation of more than 2-3 parameters. Judged by the LOOIC information criterion, the unstructured model and the model with a parameter for the susceptibility of children performed best, while the model with full age dependence was overparameterized. Estimated susceptibility of children in the model with variable susceptibility is 0.67 (95%CI: 0.40-1.1), and this is a robust finding in all models that include age-dependent susceptibility.

**Table 3.** Estimation of household transmission rates. Parameter estimates are represented by posterior medians and 95% credible intervals.

|  | Transmission rate<sup>1</sup> | Transmissibility of children<sup>2</sup> | Transmissibility of adolescents<sup>2</sup> | Susceptibility of children<sup>2</sup> | Susceptibility of adolescents<sup>2</sup> | LOOIC |
| --- | --- | --- | --- | --- | --- | --- |
| Unstructured | 1.2<br>(0.94, 1.5) | - | - | - | - | 185.6 |
| Age-dependent transmissibility | 1.0<br>(0.72, 1.4) | 0.73<br>(0.042, 2.6) | 2.7<br>(0.98, 5.6) | - | - | 186.4 |
| Age-dependent susceptibility | 1.4<br>(0.96, 2.1) | - | - | 0.65<br>(0.35, 1.1) | 0.89<br>(0.47, 1.6) | 186.5 |
| Full age-dependence | 1.2<br>(0.79, 2.0) | 0.77<br>(0.047, 2.6) | 2.3<br>(0.79, 5.2) | 0.74<br>(0.39, 1.4) | 0.93<br>(0.51, 1.7) | 189.1 |
| Susceptibility of children | 1.4<br>(1.0, 1.8) | - | - | 0.67<br>(0.40, 1.1) | - | 184.6 |

<sup>1</sup>Refers to the reference class, viz. adults.
<sup>2</sup>Transmissibility and susceptibility are relative to adults, and include intrinsic differences between groups (e.g., differences in viral loads) and varying rates at which contacts are made.

## Discussion

Estimated household secondary attack rates in our study were high (43%) and substantially higher than reported in earlier studies (reviewed in [8]). Transmission model analyses corroborated this finding and in addition revealed that children below the age of 12 had reduced susceptibility compared to adolescents and adults. Household size, severity of infection of the index case did not have a significant impact on household SAR or transmission in households.

Our study confirmed that the SAR for SARS-CoV-2 is higher than the SARs of related emerging coronaviruses such as SARS-CoV (6.0%) or MERS-CoV (3.5%) [11]. High estimated SARs are also in line with observations from surveillance data and cluster reports that the household is the most frequently reported setting of infection [26]. Our study differs from earlier household studies for SAR-CoV-2 in that we observed substantially higher SARs [11, 27]. In fact, only a few studies reported estimates that were somewhat similar to our estimates (32%-38% vs 43%) [28, 29]. A systematic review showed that estimated SARs increase with frequency of testing [27], and that most of the earlier studies analyzed existing data from contact tracing procedures performed by local public health services, monitored household contacts only during quarantine without additional follow-up, or only tested symptomatic household contacts. A few studies did test all household contacts irrespective of symptoms. However, none had a follow-up of more than 4 weeks, or an increased sensitivity for case finding through assessment of multiple sample types and diagnostic methods. Thus, we believe that our estimates of SARs may be more representative of the true household SARs than those presented in earlier studies.

A systematic review based on 18 studies concluded that children are at lower risk of infection than adults, although there was substantial heterogeneity in study design and in population characteristics [30]. Potential contributing factors include age-specific differences in the balance between innate and adaptive immunity responses [31, 32], more concomitant viral infections or cross-immunity to other coronaviruses in adolescents and adults [33-35], and physiological differences in the respiratory tract of children as compared to adults [32, 36]. Irrespective of the cause, a relevant question is how infectious infected children are to other household members, especially as in some studies the severity of infection of the index case was associated with higher infectiousness [11, 27]. We did not find such evidence for lower transmissibility of children compared to adolescents and adults, but it should be noted that our study (and for that matter, most other household studies) may be underpowered to detect even moderate differences in age-specific transmissibility.

We discuss two related limitations of our analyses. For inclusion of households our study depended on the prevailing testing policies and infected population in difference age groups. This may well have resulted in a high likelihood of selecting (symptomatic) adult index cases, such that index cases may not be representative of infections in the population. Standard practice for estimating SAR partly solves this problem by taking into account all secondary infections in the household while leaving the index case out from the analyses [20]. Related to this is the fact that the index case may not always be the primary (i.e. first) case in the household [19, 20], such that standard estimates of SARs may not be indicative of transmission routes in the household. A previous household study tried to solve this issue by including index cases and excluding primary cases [37], but this introduces bias as it would artificially increase the SAR of the prevailing type of the index cases (i.e. adults). In a sensitivity analysis, we reran the analyses using logistic regression by excluding both the primary and index case, and found relatively small impact on the estimated SARs in different age groups (not shown). The transmission model analyses do not suffer from these problems, but they do require that the primary case in the household can be identified, and this is often not possible in retrospective household analyses.

## Conclusion

Our results confirm and reinforce that the household is a main source of transmission of SARS-CoV-2, and underscore the need not only for isolation of infected household members, but also for prompt and effective quarantine of household contacts.

## Data Availability

The datasets generated and/or analyzed during the current study are not publicly available, as the data contains information that could compromise privacy and consent of participants. Data are however available from the authors upon reasonable request. Details on the transmission model used for analysis will be made available on our digital repository (https://github.com/mvboven/COVID-19-FFX).

## Acknowledgements

We thank the Public Health Service Utrecht for assistance in the recruitment of households. We thank Alper Çevirgel, Anneke Westerhof, Anne-Marie van den Brandt, Anoek Backx, Bas van der Veer, Elma Smeets-Roelofs, Elsa Porter, Elske Bijvank, Fion Brouwer, Francoise van Heiningen, Gabriel Goderski, Gert-Jan Godeke, Harry van Dijken, Helma Lith, Hinke ten Hulscher, Ilse Akkerman, Ilse Schinkel, Jeroen Hoeboer, Jolanda Kool, Josine van Beek, Joyce Greeber, Kim Freriks, Lidian Izeboud, Lisa Beckers, Lisa Wijsman, Liza Tymchenko, Maarten Emmelot, Maarten Vos, Margriet Bisschoff, Marieke Hoogerwerf, Marit de Lange, Marit Middeldorp, Marjan Bogaard, Marjan Kuijer, Martien Poelen, Nening Nanlohy, Olga de Bruin, Rogier Bodewes, Ruben Wiegmans, Sakinie Misiedjan, Saskia de Goede, Sharon van den Brink, Sophie van Tol, Titia Kortbeek, and Yolanda van Weert for support during the early stages of this study, household visits, and laboratory analyses. We would like to thank Johan Reimerink in particular for his contributions to developing the serological assays.

